# Quantitative model predicts implementing school cafeteria share tables will not compromise milk safety

**DOI:** 10.1101/2025.06.11.25329416

**Authors:** Gabriella Pinto, Yiyi Li, Melissa Pflugh-Prescott, Matthew J. Stasiewicz

**Author notes:** Address of correspondence: Matthew J. Stasiewicz, 1302 W Pennsylvania Ave. Urbana, IL 61801. Department of Nutrition, School of Medicine, Case Western Reserve University, Cleveland, OH 44106.

## Abstract

School cafeteria share tables can address food waste and improve food security by allowing students to share unopened items, like milk. However, unresolved safety concerns present a barrier to recovering milk on share tables. We adapt our previous share table model to study *Listeria monocytogenes* in pasteurized milk, assuming a concentration distribution that reflects the realistically low prevalence of the pathogen. Student sharing behavior was simulated for 50 years of school weeks (5 days/week over 37 weeks/year). Milk safety is assessed by quantifying (i) time to 1-Log_10_ *L. monocytogenes* growth, (ii) *L. monocytogenes* concentration at consumption, and (iii) listeriosis risk. We compare these measures across 22 what-if scenarios to identify potential risk factors and mitigation strategies. Under the baseline scenario (with no share table temperature management), *L. monocytogenes* increases 1-Log_10_ after one reservice (after Day 2). When there is share table temperature management, improved overnight refrigeration, or shorter services, *L. monocytogenes* did not increase by 1-Log_10_ until after two days of reservice (after Day 3). Under excessive time-temperature abuse – inadequate overnight refrigeration or long services – *L. monocytogenes* increases 1-Log_10_ before the first day of reservice (before Day 2). Comparing the baseline share table and no share table scenarios, *L. monocytogenes* concentration at consumption does not meaningfully differ. Importantly, *L. monocytogenes* concentration at consumption never exceeds 100 CFU/ml except under the longest (266 min) service scenario, for only 0.0006% of milks (11/1,794,887). The mean probability of illness per serving (P_illness_) was low across all scenarios. Comparing the baseline share table and no share table scenarios, P_illness_ was 3.32X10^-13^ and 2.72X10^-13^, respectively, translating to one listeriosis illness in every 2,100 and 3,000 years across all public schools in the United States. These results demonstrate the trivial listeriosis risk associated with recovering milk on share tables and identify practical strategies to extend reservice. Our work can support evidence-informed share table policies and assist school nutrition staff in conversations with health departments during variance processes.

**Interpretive Summary:** School nutrition programs in the United States have implemented share tables to reduce plate waste in cafeterias and improve nutrition security for students. However, concerns persist over whether the safety of Time/Temperature Controlled for Safety items, particularly milk, is compromised by implementing share tables. Thus, there is a research need to evaluate whether implementing share tables under realistic conditions would introduce food safety risk. Data generated here could support evidence-informed share table policies and assist school nutrition staff in conversations with health departments during variance processes.

## Introduction

In the 2023-2024 school year, 7 billion breakfasts and lunches were served through both the United States Department of Agriculture’s (USDA) School Breakfast Program (SBP) and National School Lunch Program (NSLP), respectively (Jones and Toosi, 2024), providing nutrition to nearly 30 million children across the United States each day (School Nutrition Association (SNA), 2024). Plate waste studies in K-12 schools have highlighted the serious waste, nutrition loss, and negative environmental impacts from discarding food and beverage items (Marlette et al., 2005; Carr and Kranz, 2012; Smith and Cunningham-Sabo, 2014; Blondin et al., 2017; World Wildlife Fund, 2019; García-Herrero et al., 2021). Notably, a large study across 8 U.S. states, including 46 schools, estimated that each school produced approximately 39 pounds of food waste per student per year, and that food waste could be costing up to $1.7 billion across all schools participating in the NSLP every school year (World Wildlife Fund, 2019). Milk was found to be one of the most wasted items in K-12 schools, specifically elementary schools, with nearly 38 cartons of milk wasted per student per year (World Wildlife Fund, 2019).

Share tables are locations in school cafeterias where whole or unopened food and beverage items can be returned (USDA, 2016) and are a low-cost strategy that can mutually benefit school nutrition programs and the children they serve. Specifically, reducing the amount of food and beverage items discarded in schools can reduce food waste and spending for school nutrition programs, which in turn can increase nutrition security for students by offering shared food and beverage items at no additional cost.

While share tables have been utilized across the U.S., there is no consistent approach that has been adopted for developing policy. The USDA provides guidance for implementing share tables; however, state or local health departments ultimately mandate which food and beverage items are allowed to be shared, resulting in variations in policy. A recent qualitative policy analysis by Prescott et al. (2020) found that 27 states had a state-level share table policy document, and 24 contained specific guidance on which food and beverage items could be shared. Importantly, nine of these policies were found to be restrictive of time/temperature controlled for safety (TCS) foods, including milk.

Previously, our work sought to demonstrate the impact on milk quality in a school cafeteria with share tables. We showed that milk spoilage as a result of contamination with a fast-growing psychrotroph, *Pseudomonas poae*, was primarily a result of overnight refrigeration (Pinto et al., 2024), and rare, even when considering worst-case conditions such as extreme refrigeration temperature, longer meal service lengths, and increased ambient temperature of cafeterias (Corea et al., 2024). However, stakeholder concerns around milk safety persist, presenting a barrier to sharing milk in school cafeterias. Here, we adapt our previous quantitative school cafeteria share table model to study *Listeria monocytogenes*, the primary foodborne pathogen of concern, in milk. Thus, the goal of this work was to assess the impact of implementing share tables on milk safety by quantifying the (i) time for a 1-Log_10_(CFU/ml) change in *L. monocytogenes* concentration in consumed milk, (ii) the *L. monocytogenes* concentration at consumption (and whether this exceeded a 100 CFU/ml threshold for concern), and (iii) the risk of listeriosis given the various realistic cafeteria conditions simulated.

## Methods

### Share Table Model Framework

Previously, we developed and adapted a quantitative share table model in R to estimate the growth of a fast-growing, psychrotrophic spoilage organism, *Pseudomonas poae*, under the time and temperature abuse expected on share tables. The time and temperature profiles were predicted for 25 realistic scenarios representing various conditions of share tables and cafeterias. The model and time-temperature profile development are described extensively in Pinto et al. (2024) and Corea et al. (2024). Briefly, the share table model simulates milk cartons and their residence time in a school cafeteria system during 5-day school weeks over a typical 37-week school year for each of 25 scenarios [see Table 1 of Corea et al. (2024)]. This is repeated 50 times to simulate 50 years of cafeteria services. In this study, we adapted the quantitative model in R 4.1.1 (R Core Team, 2025) to estimate the growth of *L. monocytogenes* for 22 scenarios proposed by Corea et al. (2024), manipulating variables such as the length of meal services (i.e., bell schedules), share table storage methods, ambient temperature of a cafeteria, and cold storage temperature, as well as a scenario with no share table. We excluded scenarios that manipulated initial contamination, as there is considerably more available data regarding the expected levels of *P. poae* from post-pasteurization contamination compared to *L. monocytogenes*.

**Table 1:** *Listeria monocytogenes* concentration model input parameters.

| Parameter Name (symbol) | Value | Units | Source | Justification/Notes |
| --- | --- | --- | --- | --- |
| *Microbial Concentration Parameters* |  |  |  |  |
| *L. monocytogenes* contamination in positive milk sample ($LM_{+}$) | -1.39 | $\text{Log}_{10}$ CFU/ml | Frye and Donnelly (2005) | 1 of 1,846 positive in skim milk, <0.3MPN/g LOD for MPN |
| Prevalence of *L. monocytogenes* positive milk ($\text{Prev}_{LM}$) | 0.05 | % | Frye and Donnelly (2005) | 1 of 1,846 samples tested positive in nonfat milk |
| *L. monocytogenes* contamination distribution ($LM_{\text{initial}}$) | Lognormal (-3.99, 0.8) | $\text{Log}_{10}$ CFU/ml | Calculated from Frye and Donnelly (2005) and Microbiological Specifications for Foods and International Commission on Microbiological Specifications for Foods (ICMSF) (2018) | |
| *Microbial Growth Model Parameters* |  |  |  |  |
| Lag time ($\lambda$) | 0 | h | Assumed value | |
| Maximum population density ($N_{\text{max}}$) | 8.5 | $\text{Log}_{10}$ CFU/ml | Koutsoumanis et al. (2010) | |
| Maximum growth rate in milk at 5°C ($\mu_{\text{max}, 5^{\circ}\text{C}}$) | 0.0181 | $\text{Log}_{10}$ CFU/h | WHO and FAO (2004) | Maximum value of the uniform distribution, converted from $\text{Log}_{10}$ CFU/d [Uniform(0.092, 0.434)] |
| Minimum growth temperature ($T_{\text{min}}$) | -2 | °C | Ramos et al. (2023) | Lowest theoretical growth temperature |
| *Dose-response Parameters* |  |  |  |  |
| Parameter r for dose response (r) | Lognormal (-12.582, 0.836) | unitless | Pouillot et al. (2024) | Selected for a virulent strain of *L. monocytogenes* in 5-14-year-old females to represent the population of primary school-aged children in our study |
| Serving size (SS) | 236 | ml | Pinto et al. (2024) |  |
| Dose (D) | $LM_{\text{Final}} \times SS$ | CFU | Calculated | |
| Number of public schools in the United States ($N_{\text{schools}}$) | 98,577 | unitless | National Center for Education Statistics (2023) | |

The cafeteria system is divided into three defined sections to simulate students interacting with meal service lines, share tables, and storage of milk cartons. (i) The meal service period, where milk cartons are served and selected, and are subsequently consumed or discarded (with or without sharing). If the milk carton is selected, it proceeds to the “sit-down” step. If it is not selected, it remains available for selection on the service line. At the sit-down step, a defined probability of consumption drives the initial “decision” to consume, share, or discard the milk carton. If the milk is consumed or discarded, the residence time in the system is recorded at the time of consumption or discarding. If the milk is shared, the selection and sit-down steps are repeated until the milk is consumed or discarded. (ii) The “break” (elapsed time) between meal service periods, in which there is no interaction between students and milk cartons. During this time, milk cartons functionally remain on share tables, aside from the one scenario where intermediate refrigeration is implemented. (iii) The overnight storage period, where milk cartons are stored under refrigeration conditions until the beginning of the next day of service. All code for the model and instructions for running it can be found on GitHub (https://github.com/foodsafetylab/Pinto-2025-Milk-Lmo).

### Hazard Identification

Several microorganisms of public health concern have been identified in raw fluid milk, particularly *Listeria monocytogenes*, *Campylobacter* spp., *Salmonella* spp., *Brucella* spp., *Yersinia enterocolitica*, pathogenic *Escherichia coli*, and *Cryptosporidium* (Schiemann, 1987; CDC, 2025). All fluid milk offered through school nutrition programs must be pasteurized to destroy microorganisms of public health concern, rendering it acceptable for sale, provided all other microbiological criteria defined by the Pasteurized Milk Ordinance (PMO) are met (FDA, 2019). However, post-pasteurization contamination has been previously reported, most commonly occurring through the introduction of Gram-negative psychrotolerant microorganisms, such as *Pseudomonas* spp., which can cause the premature spoilage of milk (Martin et al., 2018). *Pseudomonas* spp. and specific pathogens, like *L. monocytogenes*, are of particular concern due to their ability to form biofilms on the surfaces of dairy processing equipment (Buchanan and Bagi, 1999; Latorre et al., 2010; Puga et al., 2018). While rare, post-pasteurization contamination of fluid milk with microorganisms of public health concern, most frequently *L. monocytogenes*, has been documented in two foodborne disease outbreaks linked to the consumption of pasteurized fluid milk (Cumming et al., 2008; Hanson et al., 2019; Sebastianski et al., 2022).

*L. monocytogenes* is a psychrotrophic, Gram-positive bacterium responsible for causing listeriosis in humans (Ryser, 2011), and is of particular concern due to its relatively high case-fatality rate [approximately 20% (CDC, 2024)] and potential to cause severe complications from illness in high-risk populations. In the U.S., the Food and Drug Administration (FDA) considers *L. monocytogenes* a “zero-tolerance” organism in ready-to-eat (RTE) foods (FDA, 2017; Archer, 2018), and *L. monocytogenes* contamination is seldom reported in U.S.-pasteurized fluid milk. For example, a large-scale retail sampling activity across four cities in the United States reported that 1 of 5,519 fluid pasteurized milk samples tested positive for *L. monocytogenes* (Frye and Donnelly, 2005). While post-pasteurization contamination of fluid milk with pathogens is rare, *L. monocytogenes* and *Y. enterocolitica* can each grow well at refrigeration temperatures in pasteurized milk products (Bursová et al., 2017; Sebastianski et al., 2022), posing a higher concern than other mesophilic foodborne pathogens. However, due to the severity of listeriosis, the demonstrated ability of *L. monocytogenes* to form biofilms with spoilage bacteria, and recent epidemiological evidence of listeriosis outbreaks from consumption of pasteurized fluid milk compared to other foodborne pathogens (Latorre et al., 2010; Ryser, 2011; Sebastianski et al., 2022), we select *L. monocytogenes* as the most relevant pathogen of concern to assess whether implementing share tables would compromise the milk safety.

### Milk Safety Model and Initial Concentration

To our knowledge, there are very limited data regarding the concentration of *L. monocytogenes* in pasteurized fluid milk. In the model, the initial *L. monocytogenes* concentration in milk cartons follows a Log_10_-normal distribution [mean = −3.99 Log_10_(CFU/g) and standard deviation = 0.8 Log_10_(CFU/g); **Table 1**]. The mean *L. monocytogenes* concentration was estimated based on the prevalence data from Frye and Donnelly (2005), and the standard deviation was assumed based on the variability of *L. monocytogenes* concentration in other food products used in sampling cases published by the International Commission on Microbiological Specifications for Foods (ICMSF) (2018). Frye and Donnelly (2005) reported that one 25-gram sample of pasteurized fluid milk tested positive out of 1,846 samples (0.05% of nonfat milk samples). We assumed the positive sample, which enumerated below the limit of detection (i.e., < 0.3 MPN/g), was contaminated at one viable cell in the 25-gram sample [0.04 CFU/g; −1.39 Log_10_(CFU/g)]. Using the Goal Seek function in Microsoft Excel (Microsoft Corporation, 2024), given a standard deviation of 0.8 Log_10_(CFU/g), we estimate a mean of −3.99 Log_10_(CFU/g) would account for 0.05% of half-pints of milk containing a concentration of −1.39 Log_10_(CFU/g). The estimated mean value was validated by comparing the value generated using the *pnorm* function in R version 4.4.1 [value not shown; (R Core Team, 2025)]. An initial concentration of *L. monocytogenes* is assigned to every milk carton in the system by drawing a value from the distribution for contamination, thereby representing very low levels of contamination based on the observed prevalence of *L. monocytogenes* in pasteurized milk.

### L. monocytogenes Growth Model

To estimate *L. monocytogenes* growth as a function of time and temperature in the system, the Buchanan three-phase linear model (Buchanan et al., 1997b) was applied, using previously observed growth parameters for *L. monocytogenes*. The selected parameter values for this model are listed in **Table 1** and were assumed to represent the “worst-case” (most ideal) growing conditions for *L. monocytogenes*, including: (i) A lag time of 0 hours; (ii) A maximum growth rate (µ_max_) at 5 from WHO and FAO (2004) risk assessment of *L. monocytogenes* in RTE foods, where we assumed the maximum value of a uniform distribution for growth rate [Uniform(0.092, 0.434) Log_10_(CFU/d)] and converted to Log_10_(CFU/h), specifically 0.0181 Log_10_(CFU/h); (iii) The lowest theoretical minimum growth temperature from Ramos et al. (2023), −2; (iv) A maximum population density of 10^8.5^ CFU/ml from Koutsoumanis et al. (2010).

The model simulates internal milk temperature dynamically, with changes at 1-minute intervals. The growth rate during the exponential phase was thus adjusted for every 1-minute interval, using the growth rate equation from (WHO and FAO, 2004), Equation 1:

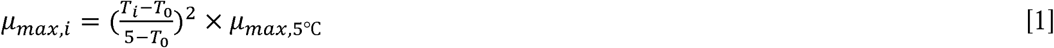

Where µ_max, I_ is the new maximum growth rate for the updated temperature in Log_10_(CFU/g h^-1^), T_i_ is the new temperature at each time interval in [, T_0_ is the theoretical minimum growth temperature, −2[, and µ_max, 5_ is the maximum growth rate, 0.0181 Log (CFU/g h^-1^).

Following this, the calculation for total growth at each interval (1-minute) was quantified as follows, Equation 2:

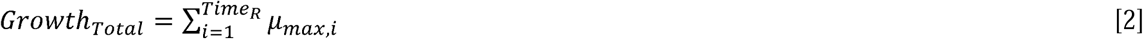

Where Growth_Total_ is the total growth in a single milk carton over its residence time in the system, and Time_R_ is the total residence time for each milk in the system.

Finally, the concentration of *L. monocytogenes* in a milk carton [in Log_10_(CFU/ml)] is estimated at the end of residence time in the system (i.e., when the milk is consumed, discarded, or donated), Equation 3:

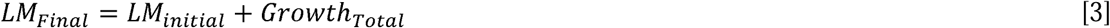

### Dose-response model

We apply an exponential dose-response function to estimate the risk of listeriosis associated with the consumption of milk in a school cafeteria under each scenario. The selected parameter values used in the dose-response model are listed in **Table 1**. The exponential dose-response model was adapted from Buchanan et al. (1997a), which is consistent with the dose-response model in the FAO/WHO (2004) *L. monocytogenes* risk assessment technical report and aligns with other recent quantitative microbial risk assessments of pasteurized milk (Ramos et al., 2023). The exponential dose-response model is shown as Equation 4:

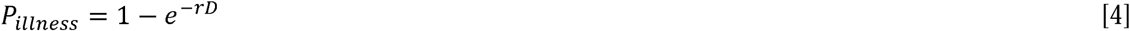

Where P_illness_ is the probability of illness (risk of listeriosis) from a single serving of a standard milk carton; *r* is the probability of developing listeriosis from digestion of one organism which is a constant that defines the shape of the dose-response curve; and *D* is the dose, number of organisms per serving, which is the antilog of LM_Final_ multiplied by the serving size [based on information reported in our previous study, 236 ml (Pinto et al., 2024)]. To better account for the variability of host susceptibility, a Log_10_-normal distribution of *r* values from Pouillot et al. (2024) was selected for a virulent strain of *L. monocytogenes* in 5 to 14-year-old females to represent the population of primary school-aged children in our study. The parameter *r* for the dose-response model was thus drawn from the distribution.

### Assessing Milk Safety in School Cafeteria Share Tables

Three metrics are used to assess milk safety based on the outputs of the share table and dose-response models: (i) the time at which a cumulative 1-Log_10_ growth of *L. monocytogenes* occurs, (ii) the *L. monocytogenes* concentration at consumption, and (iii) the listeriosis risk from consuming milk.

i. We calculate the time at which a cumulative 1-Log_10_ growth of *L. monocytogenes* is reached in each consumed milk for each scenario, and, using this information, quantify the total number of consumed milks that reached 1-Log_10_ growth on each day of meal services.
ii. We assess the concentration of *L. monocytogenes* in all simulated milks and determine whether a concentration of 100 CFU/ml is ever reached at consumption. In our model, this is considered a threshold for safety in alignment with the amended European Union Regulation No 2073/2005, which states that food business operators must demonstrate RTE foods that support the growth of *L. monocytogenes*, like milk, do not exceed 2 Log_10_(CFU/ml) of *L. monocytogenes* during product shelf life, else *L. monocytogenes* must not be detected in a 25-gram sample (European Commission, 2024).
iii. We use the calculated mean probability of illness per serving of consumed milk for each scenario to determine the risk of listeriosis. The mean value was subsequently used to determine the time that would be required for 1 listeriosis case to be observed across all schools in the United States, Equation 5:

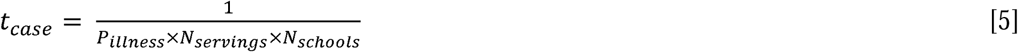

Where t_case_ represents the time elapsed in years before 1 listeriosis case is observed, P_illness_ is the mean probability of illness from consuming a single serving calculated from Equation 5, N_servings_ is the number of servings consumed per year per school simulated by the model, and N_schools_ is the number of public schools in the United States, 98,577 (National Center for Education Statistics, 2023).

## Results

### Milk is usually consumed before *L. monocytogenes* concentration increases by 1-Log_10_, which typically occurs after the second day of meal services

Results from the share table model indicate that nearly all milk consumption (99.5-100%) occurs within the first two days of service (see **Supplemental Table S1**), typically before the *L. monocytogenes* concentration increases by 1-Log_10_ under most conditions (**Table 2**). Under the baseline scenario, *L. monocytogenes* concentration does not reach a 1-Log_10_ increase until 1.62 days, after the end of meal services on Day 2 (1.09 d); where a share table with no temperature control [22.1 (72°F)] is implemented over two meal services each day (50 minutes each with a 25-minute break; 125 minutes total). Importantly, only 0.2% of total milks consumed are in the system long enough to reach this increase. *L. monocytogenes* concentration similarly does not reach a 1-Log_10_ increase until 1.37 to 2.04 days, after the end of services on Day 2 (1.09 d), under scenarios where (i) relatively mild temperature management strategies (i.e., refrigerated trays on share tables or refrigerating milk between services) are implemented, (ii) ambient temperature of the cafeteria is adjusted [18.3 (65°F), 21.1 (70°F), or 23.8 (75°F)], (iii) overnight refrigeration temperature is slightly improved [from approximately 4.4 to 4 (39.2°F)], and (iv) the duration of services remains the same as baseline, but the number of services is increased (medium services; 125 minutes, three services). Only 0.006% to 0.3% of total milks consumed are in the system long enough to reach this increase under these scenarios.

**Table 2:**
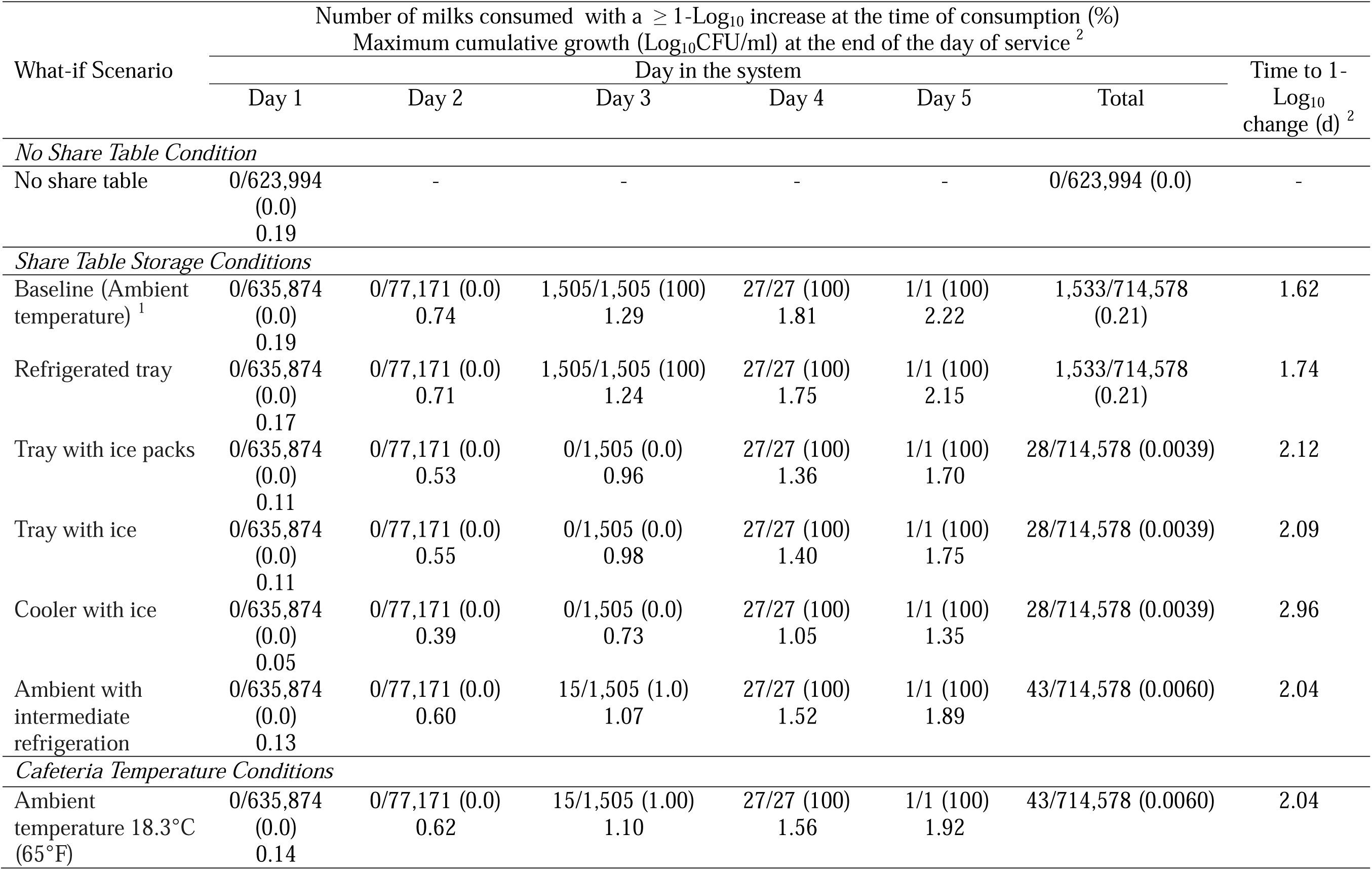
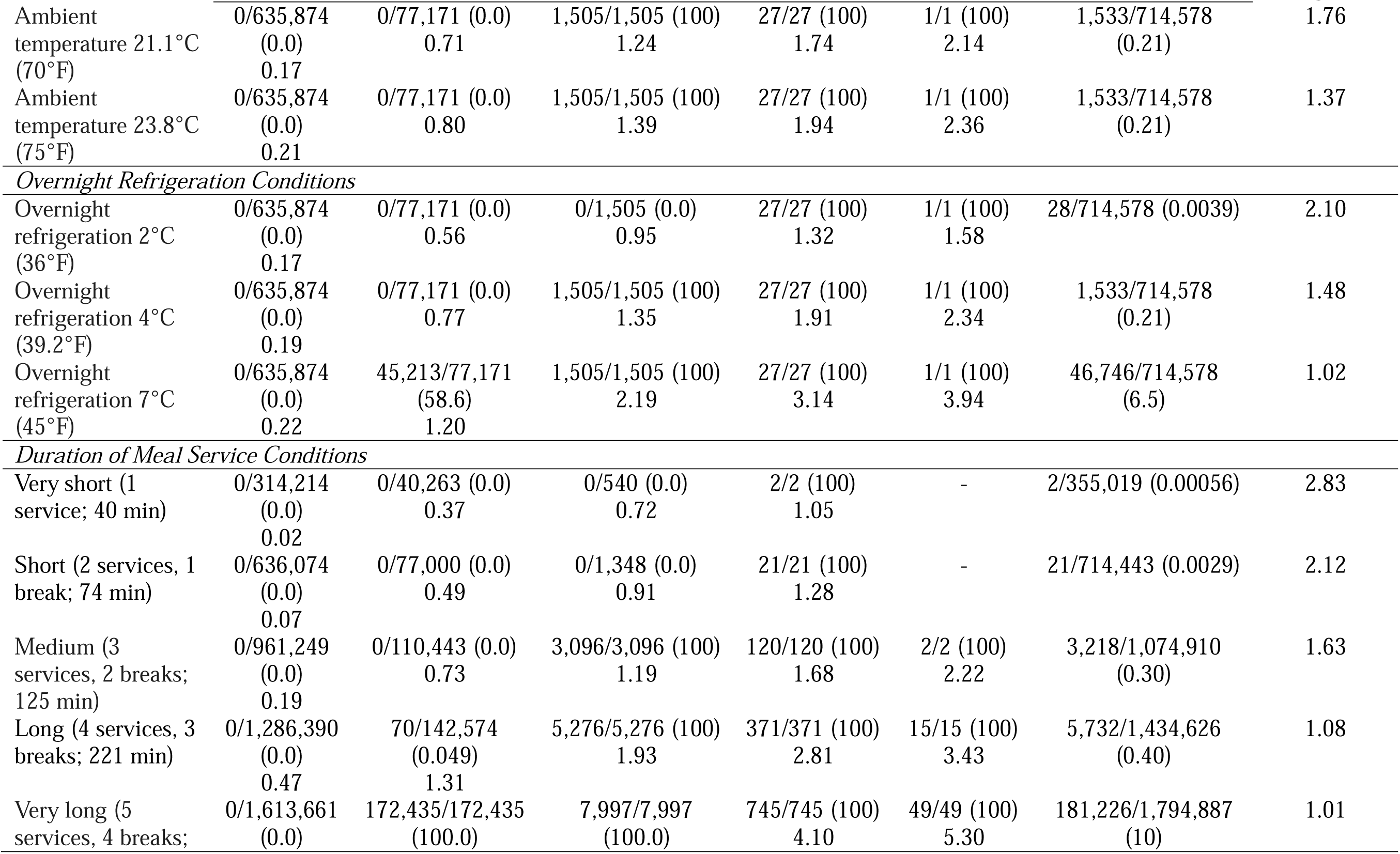
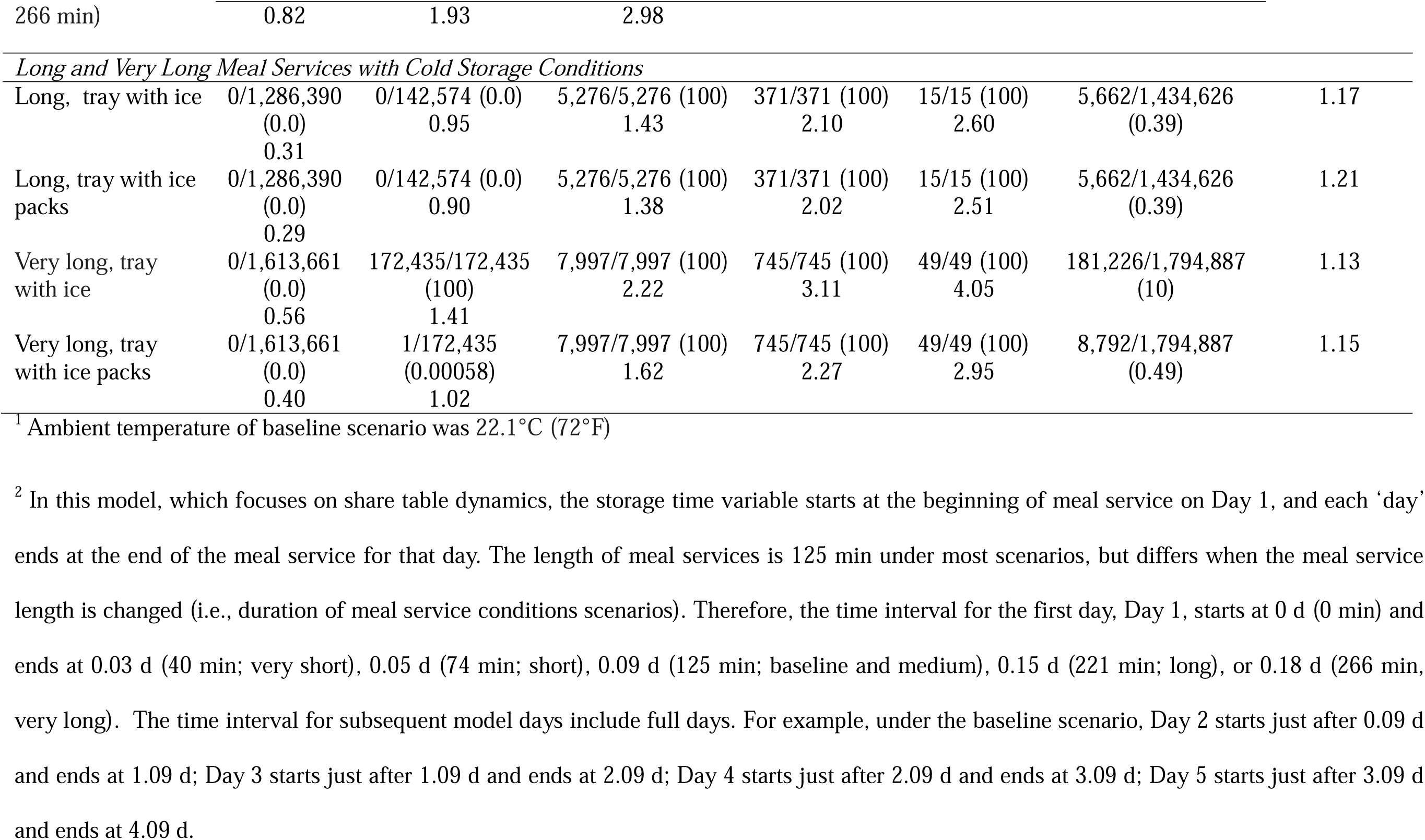
Number of consumed milks with greater than 1-Log_10_(CFU/ml) growth of *L. monocytogenes* relative to the total number of consumed milks per day of meal service, and the maximum cumulative growth [Log_10_(CFU/ml)] by the end of meal services each day.

Other conditions studied extended or reduced the amount of time to observe a 1-Log_10_ increase in *L. monocytogenes* (**Table 2**). Scenarios that extended time to a 1-Log_10_ increase (i.e., slower microbial growth) included: (i) temperature management strategies implemented on share tables (i.e., trays with ice packs, trays with ice, and coolers with ice), (ii) meaningfully lower overnight refrigeration temperature [2 (36°F)], and (iii) reducing the duration of services to very short (40 minutes; one service) or short (74 minutes; two services). For these scenarios, *L. monocytogenes* concentration does not reach a 1-Log_10_ increase until 2.10 to 2.96 days, after the end of services on Day 3 [2.03 d (very short services), 2.05 d (short services), or 2.09 d (all other scenarios with 125-min total services)]. Only 0.0006% to 0.004% of total milks consumed are in the system long enough to reach this increase under these scenarios. Overall, shorter time-temperature abuse and the addition of temperature control on shared tables and during overnight refrigeration slow the growth of *L. monocytogenes*.

Conversely, scenarios that reduced the time to a 1-Log_10_ increase (i.e., faster microbial growth) included: (i) increasing overnight refrigeration temperature [7 (45°F)], and (ii) increasing the duration of services is increased to long (221 minutes; four services) or very long (266 minutes, five services). For these scenarios, *L. monocytogenes* concentration reaches a 1-Log_10_ increase between 1.01 and 1.08 days, before the end of services on Day 2 [1.09 d (increased overnight refrigeration temperature), 1.15 d (long services), or 1.18 d (very long services)]. The increased overnight refrigeration temperature and longer time-temperature abuse each day result in between 0.4% and 10% of total milk consumed reaching a 1-Log_10_ increase. Implementing temperature management strategies on share tables (i.e., trays with ice or ice packs) under the long service scenario helps extend the amount of time until a 1-Log_10_ increase of *L. monocytogenes* concentration is reached, between 1.17 to 1.21 days, after meal services on Day 2 (1.15 days) instead of before the end of meal services on Day 2. The number of total milks that reach a 1-Log_10_ increase at consumption remains at about 0.4%. Under the very long service scenario, however, only implementing trays with ice packs on share tables helps extend the amount of time until a 1-Log_10_ increase of *L. monocytogenes*, 1.15 days, compared to 1.01 days without temperature management. While this occurs just before the end of meal services on Day 2 (1.18 days), only one milk (1/172,435; 0.0006%) reaches a 1-Log_10_ increase before the end of meal services on Day 2, compared to all milks (172,435/172,435) reaching this increase when there is no temperature management. The number of total milks that reach 1-Log_10_ increase at consumption decreases from 10% to 0.5%. Under the no share table scenario, *L. monocytogenes* never reached a 1-Log_10_ increase, as no milks were reserviced and *L. monocytogenes* does not increase by this much in 125 min.

### *L. monocytogenes* concentration in milk at the point of consumption rarely exceeded a 100 CFU/ml threshold for concern

Across all 22 what-if scenarios, the mean and maximum *L. monocytogenes* concentration in consumed milks were evaluated and are listed in **Table 3**. Despite varying conditions, the mean *L. monocytogenes* concentration at the point of consumption ranged from −3.9 to −3.5 Log_10_(CFU/ml), and the 5^th^ and 95^th^ percentiles ranged from −5.0 to −2.0 Log_10_(CFU/ml). The mean concentration at the point of consumption is generally similar to the mean of the initial contamination distribution, −3.99 Log_10_(CFU/ml). This aligns with expectations, as approximately 89.0 to 89.9% of milk is consumed on the first day of meal services, and 99.5-100% is consumed by the second day (see **Supplemental Table S1**). That is, a low median residence time permits limited growth of *L. monocytogenes*. A very small proportion of the milks remain in the system for up to five days, providing opportunities for meaningful growth of *L. monocytogenes*. To this end, we observed that the *L. monocytogenes* concentration in milk consumed after three days of reservice exceeded a 100 CFU/ml threshold for concern under the very long service scenario, reaching up to 2.8 Log_10_(CFU/ml). However, this only represented 0.0006% of the milk consumed (11/1,794,887; see **Supplemental Table S2**). Implementing temperature controls on share tables during very long services mitigated the potential for reaching this threshold, as the maximum *L. monocytogenes* concentration in any of the 1,794,887 milks consumed was reduced to 1.0 to 1.5 Log10(CFU/ml) when trays with ice packs or trays with ice were used, respectively.

**Table 3:** Mean [5^th^-95^th^ percentile] and maximum *L. monocytogenes* concentration in milk [Log_10_(CFU/ml)] at consumption across 22 what-if scenarios.

| What-if Scenario | *L. monocytogenes* concentration at consumption (Log<sub>10</sub>CFU/ml) |  |
| --- | --- | --- |
|  | Mean [5<sup>th</sup>-95<sup>th</sup> PT] | Maximum |
| *No Share Table* |  |  |
| No share table | -3.9 [-5.2, -2.6] | -0.11 |
| *Share Table with Different Storage Conditions* |  |  |
| Baseline (Ambient temperature) <sup>1</sup> | -3.9 [-5.2, -2.5] | 0.01 |
| Refrigerated tray | -3.9 [-5.2, -2.5] | -0.03 |
| Tray with ice packs | -3.9 [-5.2, -2.6] | -0.26 |
| Tray with ice | -3.9 [-5.2, -2.6] | -0.24 |
| Cooler with ice | -3.9 [-5.3, -2.6] | -0.29 |
| Ambient temperature with intermediate refrigeration | -3.9 [-5.2, -2.5] | -0.16 |
| *Cafeteria Room Temperature Conditions* |  |  |
| Ambient temperature 18.3°C (65°F) | -3.9 [-5.2, -2.5] | -0.15 |
| Ambient temperature 21.1°C (70°F) | -3.9 [-5.2, -2.5] | -0.03 |
| Ambient temperature 23.8°C (75°F) | -3.8 [-5.2, -2.5] | 0.08 |
| *Overnight Refrigeration Conditions* |  |  |
| Overnight refrigeration 2°C (36°F) | -3.9 [-5.2, -2.6] | -0.23 |
| Overnight refrigeration 4°C (39.2°F) | -3.9 [-5.2, -2.5] | 0.07 |
| Overnight refrigeration 7°C (45°F) | -3.8 [-5.2, -2.4] | 0.88 |
| *Length of Meal Service Conditions* |  |  |
| Very short (1 service; 40 min) | -3.9 [-5.3, -2.6] | -0.49 |
| Short (2 services, 1 break; 74 min) | -3.9 [-5.2, -2.6] | -0.04 |
| Medium (3 services, 2 breaks; 125 min) | -3.9 [-5.2, -2.5] | 0.20 |
| Long (4 services, 3 breaks; 221 min) | -3.7 [-5.1, -2.3] | 0.81 |
| Very long (5 services, 4 breaks; 266 min) | -3.5 [-5.0, -2.0] | 2.8 <sup>2</sup> |
| *Long and Very Long Meal Service with Cold Storage Conditions* |  |  |
| Long service, tray with ice | -3.8 [-5.2, -2.5] | 0.55 |
| Long service, tray with ice packs | -3.8 [-5.2, -2.5] | 0.53 |
| Very long service, tray with ice | -3.7 [-5.1, -2.3] | 1.5 |
| Very long service, tray with ice packs | -3.8 [-5.1, -2.4] | 1.0 |

<sup>1</sup> Ambient temperature of baseline scenario was 22.1°C (72°F)
<sup>2</sup> 11 out of 1,794,887 (0.0006%) consumed milks contain *L. monocytogenes* >2 Log<sub>10</sub>CFU/ml

### Sharing milk does not meaningfully increase *L. monocytogenes* concentration in milk during meal services

Figure 1 compares the change in *L. monocytogenes* concentration in consumed, discarded, and donated milks under the baseline (with a share table) and the no share table scenarios for the first day of meal services (Day 1). Because the majority of milk that gets consumed – 89% under the baseline scenario and 100% under the no share table scenario (**Supplemental Table S1**) – is consumed on Day 1, only these data are presented. Despite sharing status, the change in *L. monocytogenes* concentration on the first day of services is not meaningfully different in consumed, discarded, and donated milks. For the baseline scenario the median change [5^th^, 95^th^ percentiles] in *L. monocytogenes* concentration is 0.09 [0.002, 0.18] Log_10_(CFU/ml) in consumed milk, 0.09 [0.002, 0.18] Log_10_(CFU/ml) in discarded milk, and 0.08 [0.08, 0.08] Log_10_(CFU/ml) in donated milk (i.e., all milk donated at the same time, thus no variation in concentration). For the no share table scenario, the median change [5^th^, 95^th^ percentiles] in *L. monocytogenes* concentration is 0.04 [0.002, 0.18] Log_10_(CFU/ml) in consumed milk and 0.08 [0.00, 0.08] Log_10_(CFU/ml) in discarded milk. No milk is donated under the no share table scenario. While there is smaller median change in *L. monocytogenes* concentration in consumed milks on Day 1 under the no share table scenario [0.04 Log_10_(CFU/ml)] compared to the baseline share table scenario [0.09 Log_10_(CFU/ml)], the range [0 to 0.19 Log_10_(CFU/ml)] is the same under both scenarios, as milk is exposed to the same time-temperature abuse under both scenarios. The more minor median change observed can be attributed to a shorter median time in the system, which is 50 minutes when there is no share table compared to 82 minutes under the baseline system (see **Supplemental Table S3**). Shorter median time in the system is expected under the no share table scenario due to the removal of the sharing dynamic.

**Figure 1:**
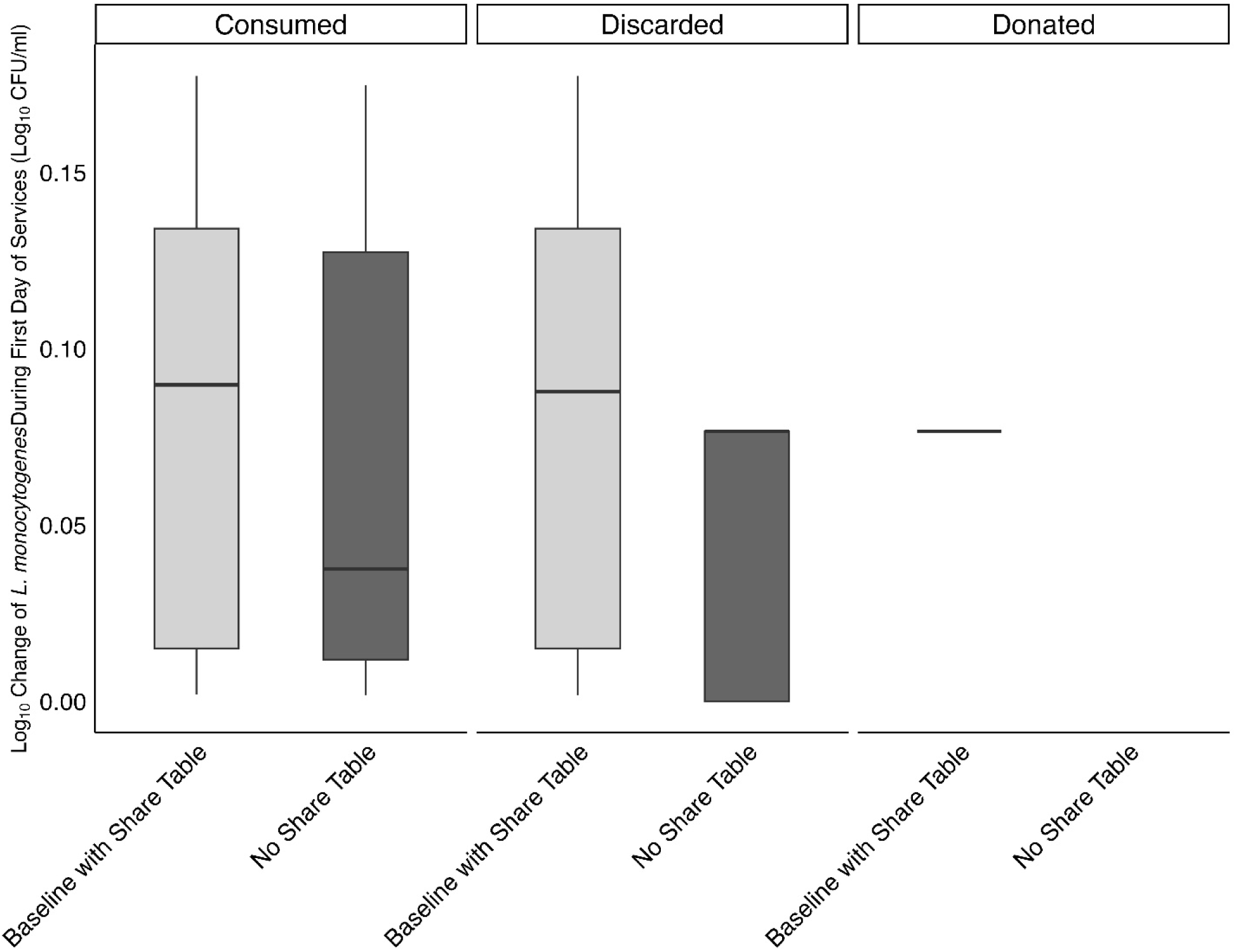
Change in *L. monocytogenes* concentration [Log_10_(CFU/ml)] in consumed, discarded, and donated milk under the baseline scenario (with a share table) and no share table scenario on the first day of services. Boxplot whiskers represent the 5^th^-95^th^ percentiles. Consumed milks are milks which students drink during the meal service. Discarded milks are milks which student throw away during the meal service. Donated milks are milks which are effectively removed and theoretically donated to another operation. Milks are not donated under the no share table scenario and can only be consumed or discarded.

### Dose-response model predicts listeriosis risk for children across public schools in the United States to be extremely low

The mean probability of illness (i) per serving, (ii) per school each year, and (iii) across all public schools each year [98,577 schools; based on the estimate from National Center for Education Statistics (2023)] among children due to the consumption of school milk generated from dose-response model are shown in **Table 4**. The mean probabilities of illness each year were extremely small across all scenarios, ranging from 3.75 × 10^-4^ to 9.72 × 10^-3^ across all public schools. The listeriosis risk remained low even when considering the most extensive time-temperature abuse scenarios such as overnight refrigeration temperature at 7 (45°F), 7.24 × 10^-4^, or very long services (t= 266 min), 9.72 × 10^-3^. To better understand risk, the number of years before a single listeriosis case would occur was estimated by taking the inverse of the mean probability of illness across all public schools each year for each scenario. Under the baseline, all storage conditions, all ambient temperatures, all overnight refrigeration, and the very short, short, and medium service length scenarios, 1,400 to 5,300 years would elapse before a single listeriosis illness would occur, given the assumptions around initial contamination of milk cartons and typical residence time in the system. Importantly, the time to a single listeriosis is extremely long across these scenarios, on the order of millennia, even when comparing the no share table scenario (3,000 years) to the baseline scenario (2,100 years). Under the more extensive time and temperature abuse scenarios (i.e., long and very long services), the time to observe a single illness ranges from 100 to 660 years. Adding temperature control methods extends the time at which this is observed to 340 to 960 years.

**Table 4:** Mean probability of illness per serving due to consumption of milk contaminated with *L. monocytogenes*, mean number of illnesses per school per year, mean number of illnesses across all public schools in the United States per year, and years to observe a single illness predicted for all school cafeteria scenarios.

| What-if scenario | Mean probability of illnesses per serving | Mean number of illnesses in one school per year<sup>2</sup> | Mean number of illnesses in all public schools per year<sup>3</sup> | Years to 1 illness ($t_{\text{cases}}$) |
| --- | --- | --- | --- | --- |
| *No share table* |  |  |  |  |
| No share table | $2.72 \times 10^{-13}$ | $3.40 \times 10^{-9}$ | $3.35 \times 10^{-4}$ | 3,000 |
| *Share Table with different storage conditions* |  |  |  |  |
| Baseline (Ambient temperature)<sup>1</sup> | $3.32 \times 10^{-13}$ | $4.74 \times 10^{-9}$ | $4.68 \times 10^{-4}$ | 2,100 |
| Refrigerated tray | $3.24 \times 10^{-13}$ | $4.63 \times 10^{-9}$ | $4.57 \times 10^{-4}$ | 2,200 |
| Tray with ice packs | $2.89 \times 10^{-13}$ | $4.13 \times 10^{-9}$ | $4.07 \times 10^{-4}$ | 2,500 |
| Tray with ice | $2.92 \times 10^{-13}$ | $4.17 \times 10^{-9}$ | $4.12 \times 10^{-4}$ | 2,400 |
| Cooler with ice | $2.66 \times 10^{-13}$ | $3.80 \times 10^{-9}$ | $3.75 \times 10^{-4}$ | 2,700 |
| Ambient with intermediate refrigeration | $3.04 \times 10^{-13}$ | $4.35 \times 10^{-9}$ | $4.29 \times 10^{-4}$ | 2,300 |
| *Cafeteria Room Temperature Conditions* |  |  |  |  |
| Ambient temperature 18.3°C (65°F) | $3.07 \times 10^{-13}$ | $4.38 \times 10^{-9}$ | $4.32 \times 10^{-4}$ | 2,300 |
| Ambient temperature 21.1°C (70°F) | $3.25 \times 10^{-13}$ | $4.64 \times 10^{-9}$ | $4.57 \times 10^{-4}$ | 2,200 |
| Ambient temperature 23.8°C (75°F) | $3.46 \times 10^{-13}$ | $4.94 \times 10^{-9}$ | $4.87 \times 10^{-4}$ | 2,100 |
| *Overnight Refrigeration Temperature Conditions* |  |  |  |  |
| Overnight refrigeration 2°C (36°F) | $2.96 \times 10^{-13}$ | $4.23 \times 10^{-9}$ | $4.17 \times 10^{-4}$ | 2,400 |
| Overnight refrigeration 4°C (39.2°F) | $3.37 \times 10^{-13}$ | $4.82 \times 10^{-9}$ | $4.75 \times 10^{-4}$ | 2,100 |
| Overnight refrigeration 7°C (45°F) | $5.14 \times 10^{-13}$ | $7.34 \times 10^{-9}$ | $7.24 \times 10^{-4}$ | 1,400 |
| *Meal Service Length Conditions* |  |  |  |  |
| Very short (1 service; 40 min) | $2.69 \times 10^{-13}$ | $1.91 \times 10^{-9}$ | $1.88 \times 10^{-4}$ | 5,300 |
| Short (2 services, 1 break; 74 min) | $2.92 \times 10^{-13}$ | $4.18 \times 10^{-9}$ | $4.12 \times 10^{-4}$ | 2,400 |
| Medium (3 services, 2 breaks; 125 min) | $3.45 \times 10^{-13}$ | $7.41 \times 10^{-9}$ | $7.31 \times 10^{-4}$ | 1,400 |
| Long (4 services, 3 breaks; 221 min) | $5.34 \times 10^{-13}$ | $1.53 \times 10^{-8}$ | $1.51 \times 10^{-3}$ | 660 |
| Very long (5 services, | $2.75 \times 10^{-12}$ | $9.86 \times 10^{-8}$ | $9.72 \times 10^{-3}$ | 100 |

| What-if scenario | Mean probability of illnesses per serving | Mean number of illnesses in one school per year <sup>2</sup> | Mean number of illnesses in all public schools per year <sup>3</sup> | Years to 1 illness ($t_{cases}$) |
| --- | --- | --- | --- | --- |
| 4 breaks; 266 min) |  |  |  |  |
| *Long and Very Long Meal Service with Cold Storage Conditions* |  |  |  |  |
| Long with tray with ice | $3.81 \times 10^{-13}$ | $1.09 \times 10^{-8}$ | $1.08 \times 10^{-3}$ | 930 |
| Long with tray with ice packs | $3.69 \times 10^{-13}$ | $1.06 \times 10^{-8}$ | $1.04 \times 10^{-3}$ | 960 |
| Very long with tray with ice | $8.43 \times 10^{-13}$ | $3.03 \times 10^{-8}$ | $2.98 \times 10^{-3}$ | 340 |
| Very long with tray with ice packs | $4.77 \times 10^{-13}$ | $1.71 \times 10^{-8}$ | $1.69 \times 10^{-3}$ | 600 |

<sup>1</sup> Ambient temperature of baseline scenario was 22.1°C (72°F)
<sup>2</sup> Number of servings varies in several scenarios: no share table scenario, very short, short, medium, long, and very long meal service length scenarios.
<sup>3</sup> Calculated using number of public schools in 2020–21 (National Center for Education Statistics, 2023)

## Discussion

The present study was conducted to assess whether sharing milk in K-12 schools via share tables compromises milk safety, and to evaluate which conditions, if any, increase or decrease listeriosis risk. We adapted our previous model for student sharing of milk cartons in a school cafeteria to study the changes in *L. monocytogenes* in pasteurized milk over 50 years of school weeks (37 school weeks/ yr, 5 d/ school week). To assess milk safety, we evaluated: (i) time for a 1-Log_10_(CFU/ml) change in *L. monocytogenes* concentration in consumed milk, (ii) the *L. monocytogenes* concentration at consumption (and whether this exceeded a 100 CFU/ml threshold for concern), and (iii) the risk of listeriosis given the various realistic cafeteria conditions simulated. We additionally compared the changes to *L. monocytogenes* concentration under the baseline share table and no share table scenarios.

While growth of *L. monocytogenes* in school milk on share tables differs across the 22 what-if scenarios evaluated – cafeteria (length of services, ambient temperature), share table (storage), and refrigeration (overnight) conditions – there is an extremely small, predicted risk of developing listeriosis from consuming milk across all scenarios. Importantly, the changes in *L. monocytogenes* concentration in milk and risk of developing listeriosis do not differ meaningfully between a system with and without a share table. The small risk of listeriosis associated with consuming milk in schools can be attributed to the low levels of contamination at consumption, on average, −3.5 to −3.9 Log_10_(CFU/ml) across all scenarios, and low median residence time in the system due to most milk consumption occurring within the first two days of services.

There is limited directly comparable scenario analysis of *L. monocytogenes* growth in pasteurized milk in the United States, as *L. monocytogenes* is considered a zero-tolerance organism. Previous quantitative microbial risk assessments (QMRAs) of pasteurized milk have varying assumptions around conditions (i.e., time and temperature), microbial growth parameters, and dose-response parameters for *L. monocytogenes*, complicating these comparisons. However, despite different conditions being modeled for *L. monocytogenes* in pasteurized milk under dynamic conditions, QMRAs have consistently shown that pasteurized milk rarely reaches concerning levels or poses a listeriosis risk to consumers (Koutsoumanis et al., 2010; Abe et al., 2023; Ramos et al., 2023). Koutsoumanis et al. (2010) developed a probabilistic model simulating *L. monocytogenes* growth during the distribution, retail storage, and domestic storage of pasteurized milk in Greece. The authors found that pasteurized milk contaminated with −2.0 or −3.0 Log_10_(CFU/ml) of *L. monocytogenes* at packaging, which is within three standard deviations of the distribution used here (**Table 1**), had a low probability of exceeding the EU 2073/2005 safety threshold of 100 CFU/ml (European Commission, 2024), 0.45% and 0.14%, respectively. Their sensitivity analysis revealed the domestic storage time, storing milk on the door shelf, and domestic storage temperature were the most important drivers of *L. monocytogenes* growth; where milk is stored at 4.98 ± 2.90 (mean ± standard deviation) for up to 96 hours in retail stores and 8.40 ± 3.00 (on the door shelf) for up to 120 hours in domestic storage.

Abe et al. (2023) and Ramos et al. (2023) both modeled *L. monocytogenes* growth in pasteurized milk, parametrizing the entire supply chain from raw milk through to the consumption of pasteurized milk. Ramos et al. (2023) included three different initial contamination [Log_10_(CFU/ml)] scenarios in raw milk, each represented by pert distributions (minimum, most likely, maximum values): 1 Log_10_ scenario (0.1, 1, 2), 3 Log_10_ scenario (2, 3, 4), or 5 Log_10_ scenario (4, 5, 6), representing a Brazilian supply chain. The results of their dose-response model showed that the minimum number of servings of pasteurized milk that would be required for a single listeriosis case in a vulnerable population was 27,248 under the highest initial raw milk contamination scenario (5 Log_10_), when milk is processed by conventional pasteurization. Under the 1 Log_10_ and 3 Log_10_ scenarios, they found that 157,729 up to 840,336 servings would be needed before observing a single listeriosis case in the vulnerable population for conventionally pasteurized milk. Through their sensitivity analysis, they showed retail storage time was the most important driver of listeriosis risk, aligning with our results that listeriosis risk was greatest when cafeteria meal service lengths were each increased. Abe et al. (2023) included two possible raw milk scenarios: no contamination and contamination from a cumulative distribution. Dose-response models were run with a different *r* parameter for perinatal, intermediate, and elderly populations. While the results of these studies are more difficult to compare because initial contamination is assumed in raw milk instead of post-pasteurization, the sensitivity analyses in both studies also showed the importance of retail and domestic storage time and temperature for pasteurized milk on illness occurrence (Abe et al., 2023) or risk of listeriosis (Ramos et al., 2023). This generally aligns with the results of our overnight refrigeration condition scenarios, which show that refrigeration temperature influences the growth of *L. monocytogenes* and, subsequently, the risk of listeriosis for a child. Under the 2 overnight refrigeration scenario, 2,400 years would elapse before a single listeriosis illness, a 12% reduction in time compared to the baseline (2,100 years). Under the 7 refrigeration scenario, 1,400 years would elapse before a single listeriosis illness occurs, representing a 35% decrease in time compared to the baseline.

There are several strengths of our analysis, including the (i) growth parameters we assumed for *L. monocytogenes* in pasteurized milk, (ii) use of dose response parameters most relevant to school-aged children, and (iii) range of scenarios model. The growth parameters selected reflect a conservative approach, assuming worst-case *L. monocytogenes* growth: 0 hours lag time [despite other relevant risk assessments assuming lag time (Xanthiakos et al., 2006; Koutsoumanis et al., 2010)], the lowest theoretical growth temperature from Ramos et al. (2023), the greatest maximum growth rate at 5 from WHO and FAO (2004), and the greatest maximum population density from Koutsoumanis et al. (2010).

Together, these factors likely result in an overestimation of risk, capturing an even greater margin of safety for sharing milk in schools than what was estimated. The use of dose-response parameters for females 5-14 years old for a virulent strain of *L. monocytogenes* from Pouillot et al. (2024), where females were more susceptible than males, also tailors the results the our population of interest, children consuming milk in K-12 schools. The greatest limitation of our analysis is that we assume all milk cartons in a given scenario are exposed to the same time and temperature profile. Additional temperature fluctuations at different positions in the refrigerator during overnight storage or when students handle milk at tables (which might be expected) could potentially result in greater *L. monocytogenes* growth in milk cartons. However, our scenario analyses for higher overnight refrigeration temperature at 7 (45°F) and higher ambient temperature in the cafeteria [23.8°C (75°F)] effectively cover the consequences of exposing contaminated milk to higher temperatures, as demonstrated by the limited increase in listeriosis risk under these scenarios.

## Conclusion

This study provide evidence for the microbiological safety of shared milk on K-12 share tables by modelling showing an extremely small risk of developing listeriosis from consuming milk from cafeterias with share tables. *L. monocytogenes* is a zero-tolerance organism in pasteurized milk and our model predicts extremely low listeriosis risk for children across public schools in the United States. Share tables do not meaningfully increase *L. monocytogenes* concentration, nor risk, in milk during meal service. Schools with short to moderate length meal services should be able to reservice milk up to three times, where schools with long meal services (3 or more hours) should limit reservice to up to two days. These results can support conversations between schools and public health departments in the variance negotiation process for share tables, and help inform the development of practical, flexible strategies tailored to individual schools.

## Notes

This project was supported by the US Department of Agriculture (USDA) National Institute of Food and Agriculture award number: [2021-68008-34106]. Any opinions, findings, or recommendations in this paper are those of the authors and do not necessarily reflect the views of the USDA. The authors have not stated any conflicts of interest.

Supplementary materials and code can be found at: https://github.com/foodsafetylab/Pinto-2025-Milk-Lmo.

No human or animal subjects were used, so this analysis did not require approval by an Institutional Animal Care and Use Committee or Institutional Review Board. The authors have not stated any conflicts of interest.

## Data Availability

All data produced are available online at https://github.com/foodsafetylab/Pinto-2025-Milk-Lmo/tree/main

