## supplemental materials for "Quantitative model predicts implementing school cafeteria share tables will not compromise milk safety"

**Table S1:** Proportion of milks consumed (%) cumulatively across a 5-day school week. All results are summed across the 50 years of iterated school years (5 days per week; 37 weeks per school year).

| What-if Scenario | Number of Milks Consumed per day/ Total Milks Consumed (%) | | | | |
| --- | --- | --- | --- | --- | --- |
|  | Day in the System ^1^ | | | | |
|  | Day 1 | Day 2 | Day 3 | Day 4 | Day 5 |
| *No Share Table Condition* | | | | | |
| No share table | 623,994/623,994  (100) | - | - | - | - |
| *Share Table Storage Conditions* | | | | | |
| Baseline (Ambient temperature) ^2^ | 635,874/714,578 (89.0) | 713,045/714,578  (99.8) | 714, 550/714,578 (99.996) | 714,577/714,578  (99.999) | 714,578/714,578  (100) |
| Refrigerated tray | 635,874/714,578 (89.0) | 713,045/714,578  (99.8) | 714, 550/714,578 (99.996) | 714,577/714,578  (99.999) | 714,578/714,578  (100) |
| Tray with ice packs | 635,874/714,578 (89.0) | 713,045/714,578  (99.8) | 714, 550/714,578 (99.996) | 714,577/714,578  (99.999) | 714,578/714,578  (100) |
| Tray with ice | 635,874/714,578 (89.0) | 713,045/714,578  (99.8) | 714, 550/714,578 (99.996) | 714,577/714,578  (99.999) | 714,578/714,578  (100) |
| Cooler with ice | 635,874/714,578 (89.0) | 713,045/714,578  (99.8) | 714, 550/714,578 (99.996) | 714,577/714,578  (99.999) | 714,578/714,578  (100) |
| Ambient with intermediate refrigeration | 635,874/714,578 (89.0) | 713,045/714,578  (99.8) | 714, 550/714,578 (99.996) | 714,577/714,578  (99.999) | 714,578/714,578  (100) |
| *Cafeteria Temperature Conditions* | | | | | |
| Ambient temperature 18.3°C (65°F) | 635,874/714,578 (89.0) | 713,045/714,578  (99.8) | 714, 550/714,578 (99.996) | 714,577/714,578  (99.999) | 714,578/714,578  (100) |
| Ambient temperature 21.1°C (70°F) | 635,874/714,578 (89.0) | 713,045/714,578  (99.8) | 714, 550/714,578 (99.996) | 714,577/714,578  (99.999) | 714,578/714,578  (100) |
| Ambient temperature 23.8°C (75°F) | 635,874/714,578 (89.0) | 713,045/714,578  (99.8) | 714, 550/714,578 (99.996) | 714,577/714,578  (99.999) | 714,578/714,578  (100) |
| *Overnight Refrigeration Conditions* | | | | | |
| Overnight refrigeration 2°C (36°F) | 635,874/714,578 (89.0) | 713,045/714,578  (99.8) | 714, 550/714,578 (99.996) | 714,577/714,578  (99.999) | 714,578/714,578  (100) |
| Overnight refrigeration 4°C (39.2°F) | 635,874/714,578 (89.0) | 713,045/714,578  (99.8) | 714, 550/714,578 (99.996) | 714,577/714,578  (99.999) | 714,578/714,578  (100) |
| Overnight refrigeration 7°C (45°F) | 635,874/714,578 (89.0) | 713,045/714,578  (99.8) | 714, 550/714,578 (99.996) | 714,577/714,578  (99.999) | 714,578/714,578  (100) |
| *Duration of Meal Service Conditions* | | | | | |
| Very short (1 service; 40 min) | 314,214/355,019 (88.5) | 354,477/355,019 (99.8) | 355,017/355,019  (99.9) | 355,019/355,019  (100) |  |
| Short (2 services, 1 break; 74 min) | 636,074/714,443 (89.0) | 713,074/714,443 (99.8) | 714,422/714,443  (99.9) | 714,443/714,443  (100) |  |
| Medium (3 services, 2 breaks; 125 min) | 961,249/1,074,910  (89.4) | 1,071,692/1,074,910  (99.7) | 1,074,788/1,074,910  (99.989) | 1,074,908/1,074,910  (99.999) | 1,074,910/1,074,910  (100) |
| Long (4 services, 3 breaks; 221 min) | 1,286,390/1,434,626  (89.7) | 1,428,964/1,434,626  (99.6) | 1,434,240/1,434,626  (99.97) | 1,434,611/1,434,626  (99.99) | 1,434,626/1,434,626  (100) |
| Very long (5 services, 4 breaks; 266 min) | 1,613,661/1,794,887  (89.9) | 1,786,096/1,794,887  (99.5) | 1,794,093/1,794,887  (99.96) | 1,794,838/1,794,887  (99.99) | 1,794,887/1,794,887  (100) |
| *Long and Very Long Meal Services with Cold Storage Conditions* | | | | | |
| Long, tray with ice | 1,286,390/1,434,626  (89.7) | 1,428,964/1,434,626  (99.6) | 1,434,240/1,434,626  (99.97) | 1,434,611/1,434,626  (99.99) | 1,434,626/1,434,626  (100) |
| Long, tray with ice packs | 1,286,390/1,434,626  (89.7) | 1,428,964/1,434,626  (99.6) | 1,434,240/1,434,626  (99.97) | 1,434,611/1,434,626  (99.99) | 1,434,626/1,434,626  (100) |
| Very long, tray with ice | 1,613,661/1,794,887  (89.9) | 1,786,096/1,794,887  (99.5) | 1,794,093/1,794,887  (99.96) | 1,794,838/1,794,887  (99.99) | 1,794,887/1,794,887  (100) |
| Very long, tray with ice packs | 1,613,661/1,794,887  (89.9) | 1,786,096/1,794,887  (99.5) | 1,794,093/1,794,887  (99.96) | 1,794,838/1,794,887  (99.99) | 1,794,887/1,794,887  (100) |

^1^ In this model which focused on share table dynamics, the storage time variable starts at the start of meal service on Day 1 and each ‘day’ ends at the end of the meal service for that day. The length of meal services is 125 min under most scenarios, but differs when meal service length is changed (i.e. duration of meal service conditions scenarios). Therefore, the time interval for the first day, Day 1, starts at 0 d (0 min) and ends at 0.03 d (40 min; very short), 0.05 d (74 min; short), 0.09 d (125 min; baseline and medium), 0.15 d (221 min; long), or 0.18 d (266 min, very long). The time interval for subsequent model days include full days. For example, under the baseline scenario, Day 2 starts just after 0.09 d and ends at 1.09 d; Day 3 starts just after 1.09 d and ends at 2.09 d; Day 4 starts just after 2.09 d and ends at 3.09 d; Day 5 starts just after 3.09 d and ends at 4.09 d.

^2^ Ambient temperature of baseline scenario was 22.1°C (72°F)

**Table S2:** Residence time (d), initial *L. monocytogenes* concentration [Log_10_(CFU/ml)], and *L. monocytogenes* concentration [Log_10_(CFU/ml)] at the time of consumption for milks that exceeded a 100 CFU/ml threshold for concern under the very long service scenario

| Milk | Residence Time in System (d) | Initial *L. monocytogenes* concentration [Log_10_(CFU/ml)] | *L. monocytogenes* concentration at consumption [Log_10_(CFU/ml)] |
| --- | --- | --- | --- |
| 1 | 4.00 | -3.08 | 2.19 |
| 2 | 4.03 | -2.69 | 2.61 |
| 3 | 4.02 | -2.56 | 2.72 |
| 4 | 3.02 | -1.52 | 2.44 |
| 5 | 4.01 | -2.51 | 2.75 |
| 6 | 3.02 | -1.90 | 2.06 |
| 7 | 4.01 | -2.58 | 2.69 |
| 8 | 4.03 | -3.02 | 2.27 |
| 9 | 4.02 | -2.55 | 2.72 |
| 10 | 4.01 | -3.15 | 2.11 |
| 11 | 4.01 | -3.24 | 2.03 |

**Table S3:** Median change in *L. monocytogenes* concentration on the first day of services [Log_10_(CFU/ml)] and median residence time in the system for consumed, discarded, and donated milks under the baseline and no share table scenarios.

| What-if Scenario | Status of Milk | Median Change [5^th^, 95^th^ Percentiles] in *L. monocytogenes* Concentration on the first day [Log_10_(CFU/ml)] | Median Residence Time in the System (min) |
| --- | --- | --- | --- |
| Baseline | Consumed | 0.09 [0.002, 0.18] | 82 |
|  | Discarded | 0.09 [0.002, 0.18] | 81 |
|  | Donated | 0.08 [0.08, 0.08] | 75 |
| No Share Table | Consumed | 0.04 [0.002, 0.18] | 50 |
|  | Discarded | 0.08 [0.00, 0.08] | 75 |
|  | Donated | N/A | N/A |
